# The effect of prone positioning on maternal haemodynamics and fetal wellbeing in the third trimester – A primary cohort study with a scoping review

**DOI:** 10.1101/2023.06.15.23291473

**Authors:** Laura Ormesher, Jessica Catchpole, Linda Peacock, Heather Pitt, Anastasia Fabian-Hunt, Dexter Hayes, Claudia Popp, Jason M. Carson, Raoul van Loon, Lynne Warrander, Karli Büchling, Alexander E P Heazell

## Abstract

**Introduction:** Supine sleep position is associated with stillbirth, likely secondary to inferior vena cava compression, and a reduction in cardiac output (CO) and uteroplacental perfusion. Evidence for the effects of prone position in pregnancy is less clear. This study aimed to determine the effect maternal prone position on maternal haemodynamics and fetal heart rate, compared with left lateral position.

**Methods:** Twenty-one women >28 weeks’ gestation underwent non-invasive CO monitoring (Cheetah) every 5 minutes and continuous fetal heart rate monitoring (MONICA) in left lateral (20 minutes), prone (30 minutes), followed by left lateral (20 minutes). Anxiety and comfort were assessed by questionnaires. Regression analyses (adjusted for time) compared variables between positions. The information derived from the primary study was used in an existing mathematical model of maternal circulation in pregnancy, to determine whether occlusion of the inferior vena cava could account for the observed effects. In addition, a scoping review was performed to identify reported clinical, haemodynamic and fetal effects of maternal prone position; studies were included if they reported clinical outcomes or effects or maternal prone position in pregnancy. Study records were grouped by publication type for ease of data synthesis and critical analysis. Meta-analysis was performed where there were sufficient studies.

**Results:** Maternal blood pressure (BP) and total vascular resistance (TVR) were increased in prone (sBP 109 vs 104 mmHg, p=0.03; dBP 74 vs 67 mmHg, p=0.003; TVR 1302 vs 1075 dyne.s-1cm-5, p=0.03). CO was reduced in prone 5.7 vs 7.1 mL/minute, p=0.003). Fetal heart rate, variability and decelerations were unaltered. However, fetal accelerations were less common in prone position (86% vs 95%, p=0.03). Anxiety was reduced after the procedure, compared to beforehand (p=0.002), despite a marginal decline in comfort (p=0.04).The model predicted that if occlusion of the inferior vena cava occurred, the sBP, dBP and CO would generally decrease. However, the TVR remained relatively consistent, which implies that the MAP and CO decrease at a similar rate when occlusion occurs. The scoping review found that maternal and fetal outcomes from 47 included case reports of prone positioning during pregnancy were generally favourable. Meta-analysis of three prospective studies investigating maternal haemodynamic effects of prone position found an increase in sBP and maternal heart rate, but no effect on respiratory rate, oxygen saturation or baseline fetal heart rate (though there was significant heterogeneity between studies).

**Conclusion:** Prone position was associated with a reduction in CO but an uncertain effect on fetal wellbeing. The decline in CO may be due to caval compression, as supported by the computational model. Further work is needed to optimise the safety of prone positioning in pregnancy.

## Introduction

The association between maternal position and cardiac output (CO) has been known for many years (1). More recently, studies have demonstrated an association between both the position in which a mother goes to sleep (2–6), and the frequency of daytime naps (2,3) and risk of late stillbirth (after 28 weeks). This association is hypothesised to be due to frequent exposure to a supine sleeping position. When a mother lies flat there is a reduction in CO and consequent uterine blood flow, this is due to compression of the inferior vena cava by the gravid uterus (1,7). These changes are associated with alterations in fetal behaviour consistent with a reduction in fetal oxygenation (8). Whilst sleep is associated with extended periods spent in specific positions, little is known about the effect of maternal position for other purposes in late pregnancy.

One study of maternal position exposed 33 Brazilian women to supine, lateral and prone positions in a random sequence (9). To maintain a prone position, woman used a specially designed concave stretcher. This study adopted each position for 6 minutes following a 10-minute period of adjustment to the experimental surroundings. This study found no differences in maternal heart rate (HR), diastolic blood pressure (dBP), oxygen saturation (SpO_2_) or fetal HR between supine, lateral and prone positions (9). However, there was a reduction in maternal respiratory rate (RR) and systolic blood pressure (sBP) when laid prone. Nevertheless, all the women reported feeling comfortable lying flat (on a bent surface).

Another study of 65 pregnant women from Australia, 15 of whom had pre-eclampsia found that lying in a prone position for 5 minutes was associated with a 2 mmHg reduction in sBP in healthy women but a larger 6 mmHg fall in women with pre-eclampsia. this was associated with a compensatory increase in HR (10). These cardiovascular effects merit further exploration. Importantly, there have been no studies investigating a more clinically meaningful timeframe of exposure to prone position for physical therapies e.g. 30 minutes.

Clearly, maintaining a prone position for physical therapies in late pregnancy is difficult due to the gravid uterus. The Anna cushion (Supplementary Figure 1) was developed to support mothers in a prone position. The concave Anna cushion is specifically moulded from a medical grade, medium density, closed cell foam, which has been covered in a double layer of cotton lycra fabric. It is deliberately shaped to accommodate for the pregnant abdomen up until the end of pregnancy.

This study aimed to describe the cardiorespiratory effects of a mother maintaining a prone position supported by the device (Anna cushion) for a period of 30 minutes. In addition, we aimed to determine whether maintaining a prone position is associated with any effects on the fetal HR, and to determine whether using a device to support a prone position is comfortable for the mother. We used the primary data from the clinical study to inform a mathematical model to determine whether compression of the inferior vena cava could explain our observations. We also conducted a scoping review to synthesise data regarding the clinical and haemodynamic effects of maternal prone positioning. This study was based on the primary hypothesis that prone positioning, with support of the Anna cushion, would be acceptable to pregnant women and associated with a decline in maternal blood pressure (BP), compared with left lateral position.

## Material and methods

### Primary Study

This single-centre prospective observational feasibility study was conducted at the Maternal and Fetal Health Research Centre, Saint Mary’s Hospital, Manchester, UK. Recruitment (via referral) and follow-up were from 13^th^ July 2021 to 23^rd^ March 2022. Women were approached to participate in the study if they met the following inclusion criteria: aged 16 to 50 with a viable singleton pregnancy at over 28 weeks’ gestation, and ability to read written English. Exclusion criteria were: evidence of fetal compromise or existing fetal anomaly, pre-existing maternal conditions that could influence the cardiovascular system, and maternal contraindications to lying prone (such as severe pain or spinal disease). Participants were offered a small sum (£25) as thanks for their participation in the study. Any unexpected adverse outcome that resulted in deviation from the study protocol automatically prompted termination of the study and immediate clinical assessment. Eligible and willing participants gave written consent to participate in the study. Data were collected onto case report forms.

Prior to commencing the experimental protocol, ultrasound examination was performed (if one had not been performed in the preceding two weeks) and baseline haemodynamic measurements were recorded. The pre-study questionnaire included questions regarding the women’s current physical comfort, their pregnancy symptoms and treatment, and their pre-pregnancy and current sleeping positions. It then went on to measure the women’s self-reported anxiety levels using the State Trait Anxiety Inventory (STAI).

Once the required maternal and fetal monitoring equipment was attached, all women began by resting in the left lateral position for twenty minutes. Participants were then asked to lie in the prone position for a further thirty minutes, supported by the Anna cushion. They then returned to the left lateral position for a final twenty minutes.

Maternal and fetal haemodynamic variables were recorded at five-minute intervals throughout the positional sequence. Timing was monitored using a digital stopwatch. Maternal haemodynamics were measured using a Non-Invasive CO Monitor (NICOM, Cheetah), a monitor that uses bioreactance to estimate cardiovascular parameters. Maternal sBP and dBP were monitored using an electronic sphygmomanometer (Omron M3) placed on the right upper arm. SpO_2_ was measured using a digital pulse oximeter placed on the left index finger. RR was measured visually by an investigator. Fetal HR was monitored using a Monica AN24 device. Fetal behavioural state was assessed on the Monica AN24 traces by two observers blinded to maternal position during the time period covered by the recording according to the fetal behavioural states described by Pillai *et al.* (11).

The post-study questionnaire was administered upon completion of the maternal positional sequence. It included questions regarding maternal comfort acceptability of the Anna cushion and study protocol. Post-study anxiety levels were then reassessed using the state component of the STAI. Only those in the primary clinical team (HP, LP, LO, AH) had access to information that could identify individual participants.

### Primary and secondary outcomes

The primary outcome was maternal cardiac output. Secondary outcomes were maternal cardio-respiratory status (including heart rate, blood pressure, respiratory rate and oxygenation via pulse oximetry), fetal wellbeing (including baseline heart rate, variability, the presence of accelerations or decelerations, and acceptability (including maternal anxiety and comfort).

### Statistical analysis

All statistical analyses were conducted in Stata (Version 14, STATACORP, TX, USA). The alpha level for statistical significance was set at 0.05. Skewness of continuous variables was assessed using the Jarque-Bera skewness-kurtosis test and histograms. Parametric data were presented as mean ± standard deviation and non-parametric data as median (interquartile range). Categorical variables were presented as absolute frequencies (%). Comfort and state anxiety scores before and after the study protocol underwent paired analysis using the Wilcoxon signed-rank test. Multivariable regression analyses compared continuous variables between positions, having adjusted for time. Categorical variables were compared between positions using Chi-square test.

The sample size for this study was calculated to determine whether maternal CO (the primary outcome measure) decreases in the prone position compared to left lateral position. Milsom *et al.* (1) demonstrated a mean CO of 6.6L/min in left lateral position, 5.9L/min in right lateral position and 5.5L/min in a supine position, with a standard deviation of 1.0L/min. To have 80% power to detect a fall from 6.6L/min (the level reported for left lateral position) to 5.7 L/min (the midpoint between right lateral and supine positions), 20 participants would be required in each group using alpha=0.05.

### Mathematical Modelling

A mathematical model of the cardiovascular system in pregnancy was implemented to investigate the mechanical impact of inferior vena cava occlusion and the consequent effects on systemic arterial pressure and CO. The framework utilised has been previously described (12,13). The cardiovascular network model contains 513 blood vessels including: the major systemic arteries, systemic veins, pulmonary arteries, and pulmonary veins. The model also includes the major organs, including the brain, lungs, liver, kidneys, spleen, intestines, uterus, and stomach. The mathematical system of equations is solved using a sub-domain collocation scheme, which is described in Carson and Van Loon’s study (14). In order to personalise the cardiovascular model, patient measurement data is incorporated into the framework through a parameter optimisation technique. The patient sBP and dBP, HR, stroke volume (SV), and height (H) are used to estimate the total vascular resistance (TVR), total arterial compliance, and vessel lengths. When of interest, pulse wave velocity (PWV) can be utilised to estimate major vessel diameters, and the maximum and minimum uterine artery blood velocities measured from Doppler ultrasound can also be incorporated into the framework (13).

In order to investigate the impact of occlusion of the inferior vena cava, the patient measurements of sBP, dBP, CO, HR, and H from the prone position cohort were used in the parameter estimation. Initially, the model was simulated until convergence to the measurement values with no inferior vena cava occlusion present. Once convergence was achieved the optimisation algorithm was switched off, meaning the model parameters of resistance, compliance, and blood volume were kept constant in any subsequent simulations. An external pressure was then incrementally applied to the outer wall of the inferior vena cava causing various levels of occlusion. The model-predicted values for sBP, dBP, and CO were then recorded for these different percentages of inferior vena cava occlusion. The occlusion occurred in inferior vena cava III in the network, which is located just below the level of the hepatic veins. This was chosen in order to cause the minimum effective dose of occlusion and to investigate the effect. If more of the venous return pathway of the inferior vena cava was occluded, the effect on flow reduction would be even greater. It was decided to avoid compressing the abdominal aorta (AA) as well, due to observations in studies that suggest AA occlusion either rarely occurs (15), or has much less compression compared to the veins (16).

### Scoping Review and Meta-analysis

A literature search was performed (by JC, DH and AH in collaboration with an information specialist) on the 18th of June 2022 using the following primary databases: Cochrane (Database of Systematic Reviews; Central Register of Controlled Trials), Medline and Embase. A grey literature search was also carried out of the databases BASE and OpenGrey using the same search terms and criteria; the search strategies were supplemented by hand searching from reference lists. In order to broaden the scope of the results, the initial search had no exclusion criteria. The search strategy consisted of the three main concepts of pregnancy, cardiorespiratory status, and the prone position. Keywords, medical subject headings (MeSH terms) and truncations were used where applicable (a summary of the search terms is available in Supplementary file 1).

After results from each database were combined, duplicate records were removed. Screening was performed manually by a single reviewer (JC or AFH) with queries resolved by discussion with a second person (AH). Records were initially screened by title and abstract and excluded if they were not relevant to the prone position in pregnancy (for example infant prone position and SIDS). Full text-assessment was then performed on the remaining 132 records. Records were excluded by the following pre-determined criteria: lack of information or outcome data available in the text regarding the use of the prone position during pregnancy; prone position discussed but not utilised; prone position utilised in postpartum period. Study records were grouped by publication type for ease of data synthesis and critical analysis. As case reports and case series were the most common, these were presented in a summary table and assessed for methodological quality using a tool modified from Murad *et al*. (17). This tool awarded each case report a validity score, using a six-point framework based on the four following domains: selection, ascertainment, causality and reporting. For the purpose of case report evaluation, the primary outcome measures of interest were livebirth and lack of fetal distress during maternal prone positioning. Adequate length of follow-up was defined as follow-up until birth or thereafter. A case report was considered adequately detailed if it provided information on the reason for use of the prone position, the method of prone positioning, and maternal and fetal outcomes.

Where quantitative synthesis was possible, random effects meta-analysis was performed using the command *metan* in STATA (Version 14). Heterogeneity was assessed using the I^2^ statistic.

### Ethics Statement

This study was approved by the Health Research Authority (IRAS ID: 240072) and North East - Newcastle & North Tyneside 1 Research Ethics Committee (20/NE/0261). The study was registered on clinicaltrials.gov (NCT04586283, https://clinicaltrials.gov/ct2/show/NCT04586283). Initial participant enrolment was on 13/07/21.

## Results

### Primary Study

Thirty-seven women were approached to participate in the study; 21 women participated in the experimental protocol; one woman had a fetal bradycardia resulting in early termination of the protocol (Figure 1). Complete data were therefore available for 20 women. The demographic characteristics of the participants are summarised in Table 1. Mean gestation at participation was 32 weeks + 6 days. Table 2 summarises the pregnancy outcomes of the cohort.

**Figure 1:**
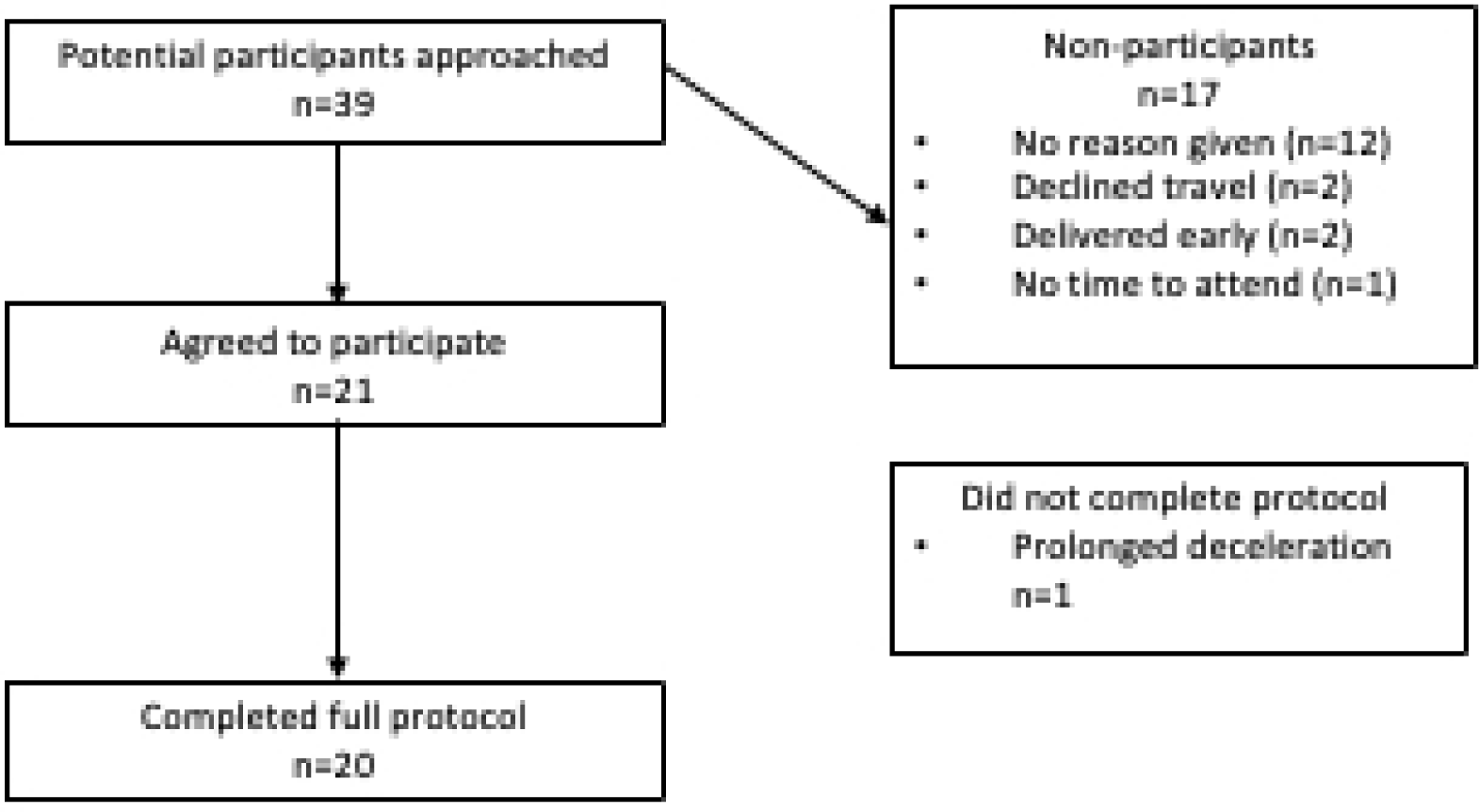
Consort diagram.

**Table 1:** Demographic characteristics of participants

| Characteristic |  | Distribution (Median (range) or n (%)) |
| --- | --- | --- |
| Maternal Age |  | 32 (26-42) |
| Ethnicity | Black African | 1 (5) |
|  | Mixed ethnicity | 1 (5) |
|  | South Asian | 2 (10) |
|  | White British | 15 (71)) |
|  | White Other | 2 (10) |
| Booking Body Mass Index (kg/m<sup>2</sup>) |  | 25.3 (19.4-36.2) |
| Gravidity |  | 2 (1-5) |
| Parity |  | 1 (0-2) |
| Booking blood pressure (mmHg) |  | 112 (98-129) / 67 (45-79) |
| Cigarette smoker |  | 2 (10) |
| Gestation at participation (weeks + days) |  | 32<sup>+0</sup> (28<sup>+6</sup>-37<sup>+5</sup>) |
| Body Mass Index at participation |  | 28.3 (22.0-37.3) |
| Maternal abdominal circumference (cm) |  | 106 (91-140) |

**Table 2:** Pregnancy outcomes of participants

| Pregnancy outcomes |  | Median (range) or N (%) |
| --- | --- | --- |
| Gestation at delivery* |  | 273 (267 – 278) |
| Birthweight |  | 3383.9 ± 602.4 |
| Birthweight centile |  | 44.9 ± 29.8 |
| Mode of delivery | Emergency Caesarean Section | 5 (24%) |
|  | Elective Caesarean Section | 3 (14%) |
|  | Instrumental Vaginal Birth | 3 (14%) |
|  | Spontaneous Vaginal Birth | 10 (48%) |
| Hypertensive disorder of pregnancy |  | 1 (5%) |
| Small for Gestational Age (<10<sup>th</sup> centile) |  | 4 (19%) |
| Fetal Growth Restriction (<3<sup>rd</sup> centile) |  | 1 (5%) |
| Perinatal mortality |  | 0 (0%) |
| Female sex |  | 11 (52%) |

There was a small reduction in the median state anxiety scores (38 vs 30, p=0.002; Figure 2) from before to after the experimental protocol. However, there was no difference in comfort scores (2 vs 2, p=0.13) before and after the protocol. There was a small reduction in anxiety score between left lateral and prone (2 vs 1, p=0.04), despite a reduction in comfort (2 vs 1, p=0.04).

**Figure 2:**
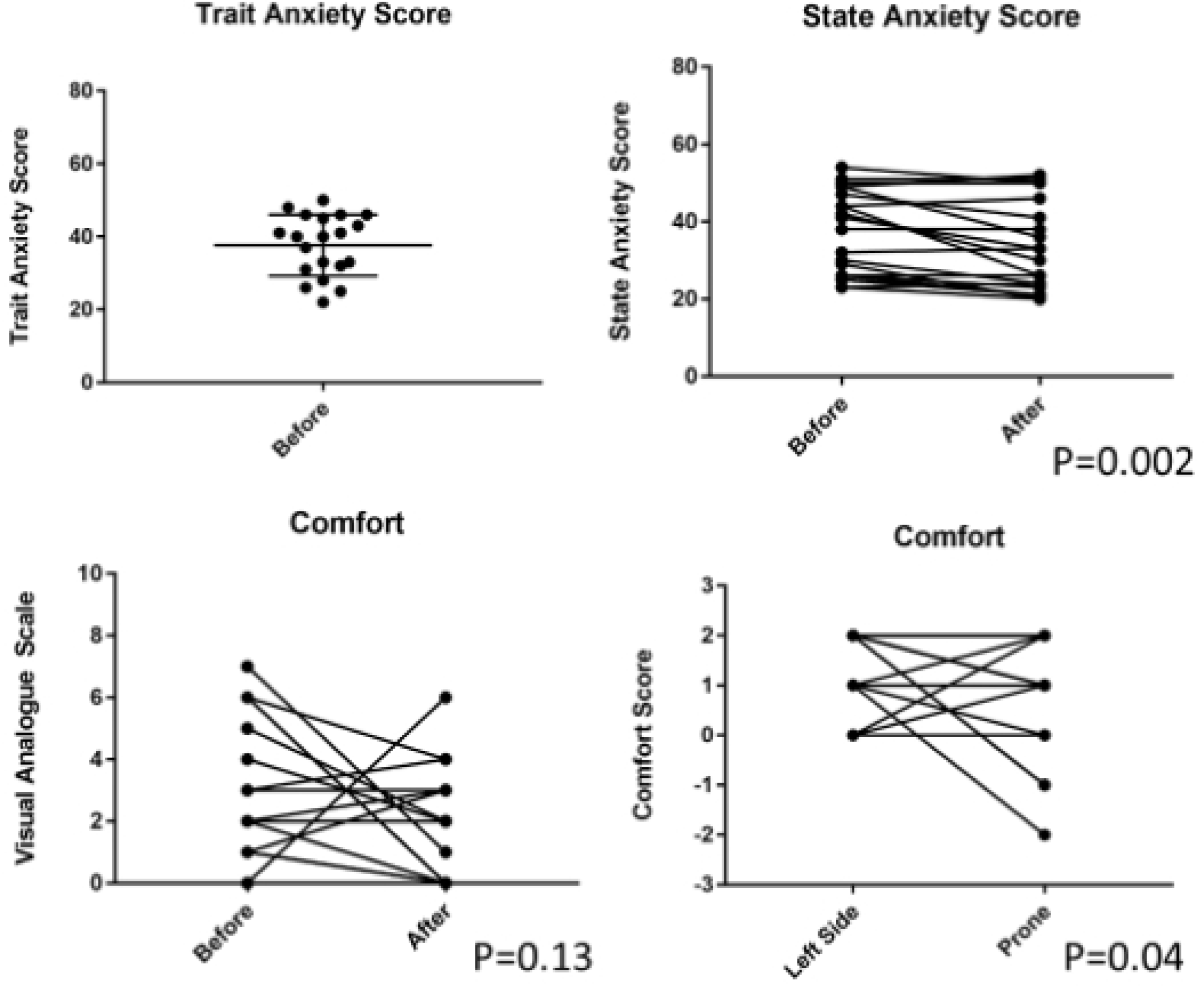
Acceptability scores before and after lying in the prone position.

Table 3 summarises maternal haemodynamics by maternal position. Maternal BP and TVR were increased in prone (sBP adjusted difference 6.0 mmHg [95% C.I. 0.6 - 11.5], p=0.03; dBP 7.0 mmHg [2.4 - 11.6], p=0.003; TVR 243.4 dyne.s^-1^cm^-5^ [29.7 - 457.0], p=0.03; Figure 3). SV and CO were reduced in prone (SV adjusted difference -20.9 mL [-32.6 to -9.1], p=0.001; CO -1.5 mL/minute [-2.5 to -0.5], p=0.003). Haemodynamic changes were consistent across the cohort (Figure 4). There were no changes in maternal RR and spO_2_ in prone position. Fetal HR, variability and decelerations were unaltered in prone position (Table 4). Fetal accelerations were less common in prone position (86% vs 95%, p=0.03), although there was no difference in fetal behavioural state (Table 4). Figure 5 illustrates a lack of temporal relationship between maternal haemodynamics and fetal HR in the woman who experienced a fetal bradycardia.

**Figure 3:**
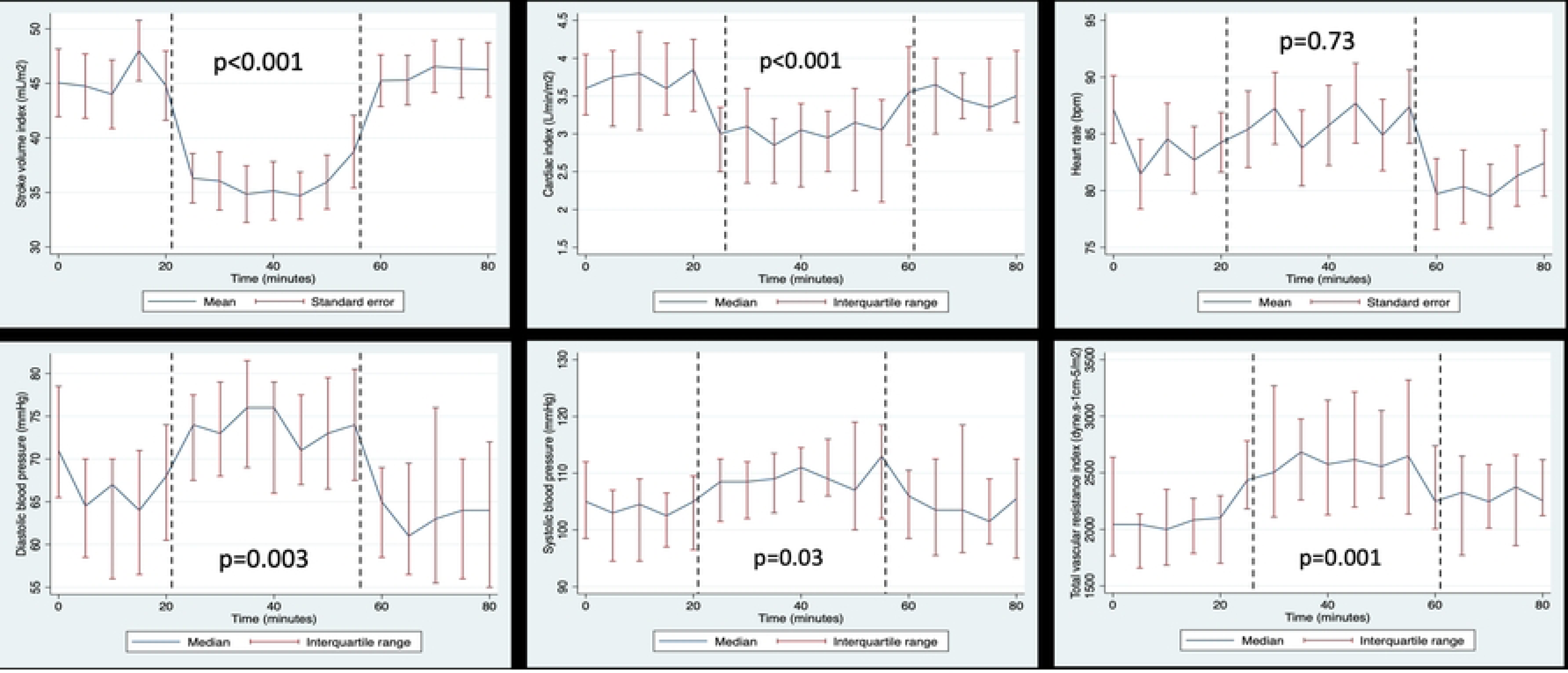
Longitudinal maternal haemodynamics by maternal position.

**Figure 4:**
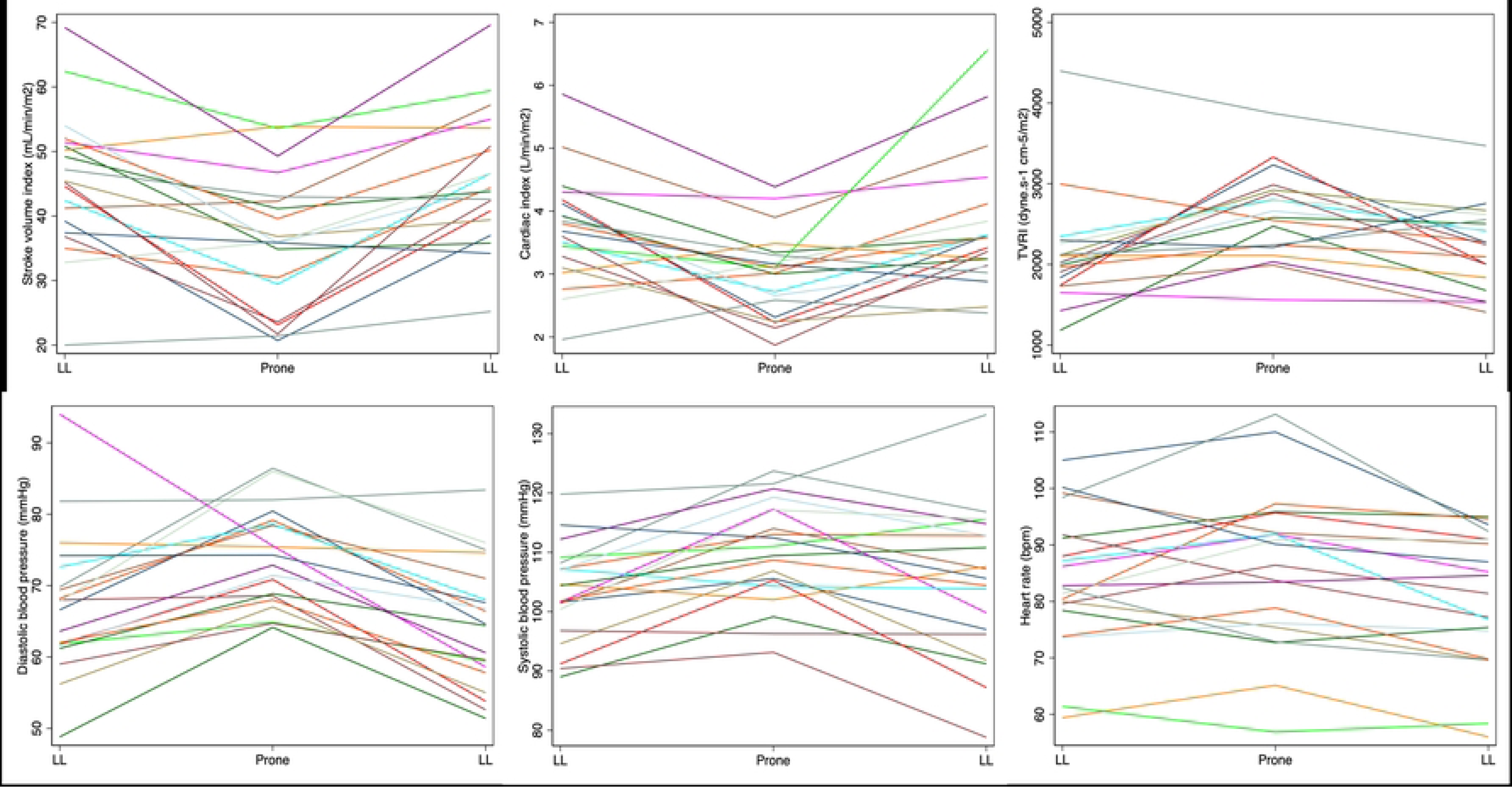
Mean positional maternal haemodynamics by individual.

**Figure 5:**
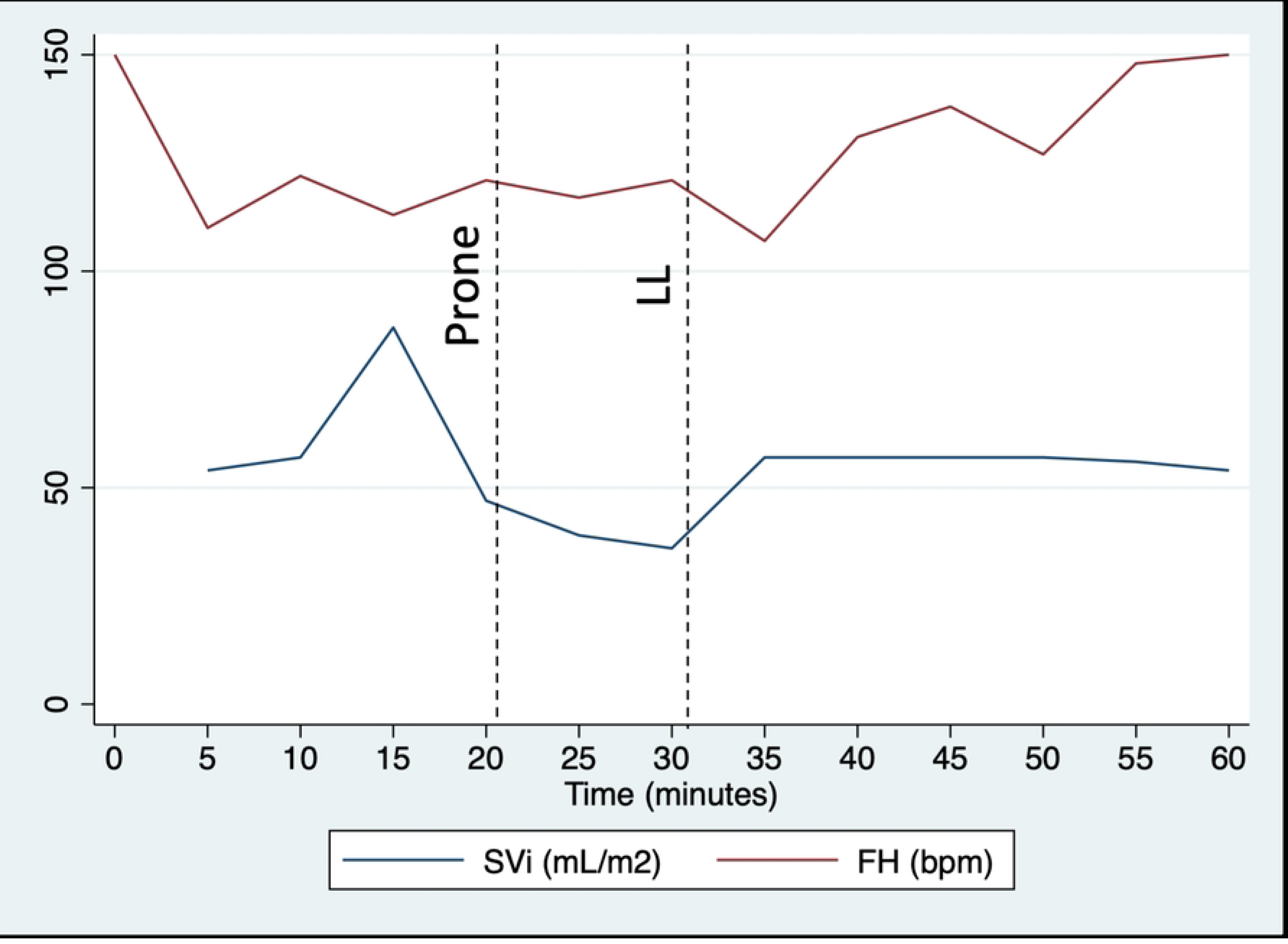
Relationship between maternal stroke volume index and fetal heart rate in the woman who required early termination of the protocol.

**Table 3.** Maternal haemodynamic and respiratory indices in left lateral and prone positions.

|  | Left lateral 1 | Prone | Left lateral 2 | Position 1 to 2 (n=20) |  | Position 1 to 3 (n=20) |  |
| --- | --- | --- | --- | --- | --- | --- | --- |
|  |  |  |  | Adjusted difference | 95% C.I. | Adjusted difference | 95% C.I. |
| **sBP (mmHg)\*** | 104 (96 - 108.5) | 109 (103 - 116.5) | 105 (96.5 - 111.5) | 6.0 | 0.6 - 11.5 | 0.2 | -9.4 - 9.9 |
| **dBp (mmHg)\*** | 67 (58.5 - 72.5) | 74 (67.5 - 79) | 63 (56 - 70) | 7.0 | 2.4 - 11.6 | -2.2 | -10.3 - 5.8 |
| **HR (bpm)** | 84.0 ± 13.1 | 86.0 ± 14.6 | 80.6 ± 13.0 | 1.1 | -5.2 - 7.5 | -5.1 | -16.3 - 6.0 |
| **RR (bpm)\*** | 16.5 (16 - 18) | 16 (16 - 18) | 17 (16 - 18) | 0.1 | -0.8 - 0.9 | 0.7 | -0.8 - 2.2 |
| **O<sub>2</sub> saturation (%)\*** | 99 (98 - 99) | 99 (98 - 99) | 99 (98 - 99) | 0.0 | -0.5 - 0.4 | -0.1 | -0.8 - 0.7 |
| **SV (mL)\*** | 88.4 (73.2 - 98.4) | 71.3 (48.8 - 84.9) | 85.4 (71.0 - 96.9) | -20.9 | -32.6 - -9.1 | -5.7 | -26.4 - 15.1 |
| **CO (L/min)\*** | 7.1 (5.7 - 8.0) | 5.7 (4.3 - 6.8) | 6.3 (5.6 - 7.6) | -1.5 | -2.5 - -0.5 | -0.5 | -2.3 - 1.2 |
| **TVR (dyne.s<sup>-1</sup>cm<sup>-5</sup>)\*** | 1075.3 (1013.2 - 1177.4) | 1302.2 (1120.6 - 1614.6) | 1228 (1063 - 1330) | 243.4 | 29.7 - 457.0 | 43.4 | -331.1 - 418.0 |
| **SVI (mL/m<sup>2</sup>)** | 45.3 ± 13.4 | 36.0 ± 11.5 | 45.9 ± 10.7 | -11.0 | -16.4 - -5.5 | -2.6 | -12.2 - 7.1 |
| **CI (L/min/m<sup>2</sup>)\*** | 3.7 (3.3 - 4.2) | 3.0 (2.3 - 3.4) | 3.5 (3.1 - 4.0) | -0.8 | -1.2 - -0.4 | -0.1 | -0.9 - 0.6 |
| **TVRI (dyne.s<sup>-1</sup>cm<sup>-5</sup>/m<sup>2</sup>)\*** | 2047.5 (1720 - 2294) | 2621.6 (2225.4 - 2921.0) | 2288 (1961 - 2640) | 516.6 | 226.5 - 808.7 | 120.1 | -401.6 - 641.8 |

Mean ± standard deviation
\*Median (range)
mmHg, millimetres of mercury; sBP, systolic blood pressure; dBp diastolic blood pressure; bpm, beats/ breaths per minute; HR, heart rate; RR, respiratory rate; SV, stroke volume; CO, cardiac output; TVR, total vascular resistance; SVI, stroke volume indexed to body surface area; CI, cardiac output indexed to body surface area; TVRI, total vascular resistance indexed to body surface area.

**Table 4.** Fetal heart rate indices and behavioural state assessment showing the mean values for baseline heart rate and variability and the proportion of cases in each fetal behavioural state per maternal position. *Fisher’s exact test Left Lateral 1 vs. Prone = 0.45, Prone vs. Left Lateral 2 = 0.55.

|  | Left lateral 1 | Prone | Left lateral 2 | Position 1 to 2 (n=20) |  | Position 1 to 3 (n=20) |  |
| --- | --- | --- | --- | --- | --- | --- | --- |
| **Fetal heart rate (bpm)** | 138.0 ± 8.2 | 139.6 ± 7.5 | 140.6 ± 9.6 | 1.5 | -5.1 - 8.0 | 1.2 | -10.3 - 12.7 |
| **Variability (bpm)\*** | 13.9 (11.2 -15.8) | 13.4 (11.1 - 15.0) | 15 (10 - 18) | -0.2 | -3.5 - 3.0 | 1.4 | -4.2 - 7.1 |
| **Fetal Behavioural State** |  |  |  |  |  |  |  |
| 1F | 1 (4.8) | 2 (9.5) | 1 (4.8) | 0.45* |  | 0.55* |  |
| 2F | 17 (81.0) | 13 (61.9) | 16 (76.2) |  |  |  |  |
| 3F | 0 (0) | 1 (4.8) | 0 (0) |  |  |  |  |
| 4F | 0 (0) | 3 (14.3) | 2 (9.5) |  |  |  |  |
| Indeterminable | 3 (14.3) | 2 (9.5) | 2 (9.5) |  |  |  |  |

bpm, beats per minute.

Change in average haemodynamic measures over time. The line represents the mean (parametric) / median (non-parametric) and the bar represents standard error (parametric) / interquartile range (non-parametric). The dashed line indicates the timing of position change. P value is derived from multivariable regression analyses with position as the independent variable, adjusted for time.

LL, left lateral; mmHg, millimetres of mercury; bpm, beats per minute.

Mean haemodynamic measures are plotted for each woman (different colour) during the three position changes.

LL, left lateral; mmHg, millimetres of mercury; bpm, beats per minute.

The dashed line indicates the timing of position change.

LL, left lateral; SVi, stroke volume indexed to body surface area; FH, fetal heart rate; bpm, beats per minute.

### Mathematical Modelling

When comparing with the patient measurements, it was postulated that some of the differences in BP and CO between the left lateral position and prone position were caused by inferior vena cava occlusion (Figure 6). The expected behaviour would be that both the mean arterial pressure (MAP) and CO would decrease, as in supine hypotensive syndrome, secondary to uterine compression of the inferior vena cava.

**Figure 6:**
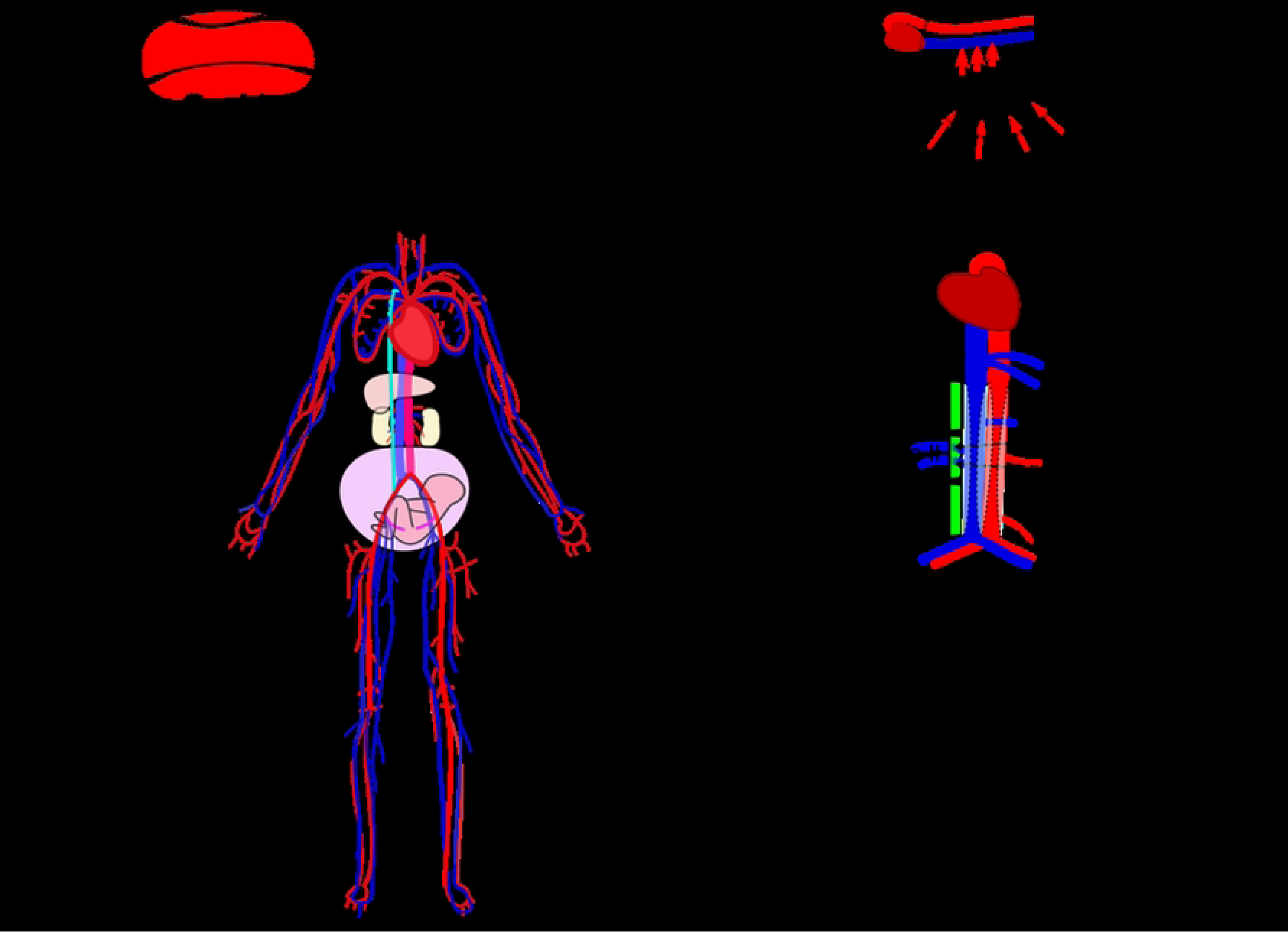
Vena cava compression during prone position (top images). A representation of the computational cardiovascular network model (left bottom) and a zoomed in version showing IVC III as the location where compression was simulated (bottom right).

Figure 7 shows the model predicted trends of sBP, dBP, CO, and TVR for the study participants. The model predicted that if occlusion of the inferior vena cava occurred, the sBP, dBP and CO would generally decrease. However, the TVR remained relatively consistent, which implies that the MAP and CO decrease at a similar rate when occlusion occurs. Our model predicted that not all participants (4 of the 21) would follow this trend.

**Figure 7:**
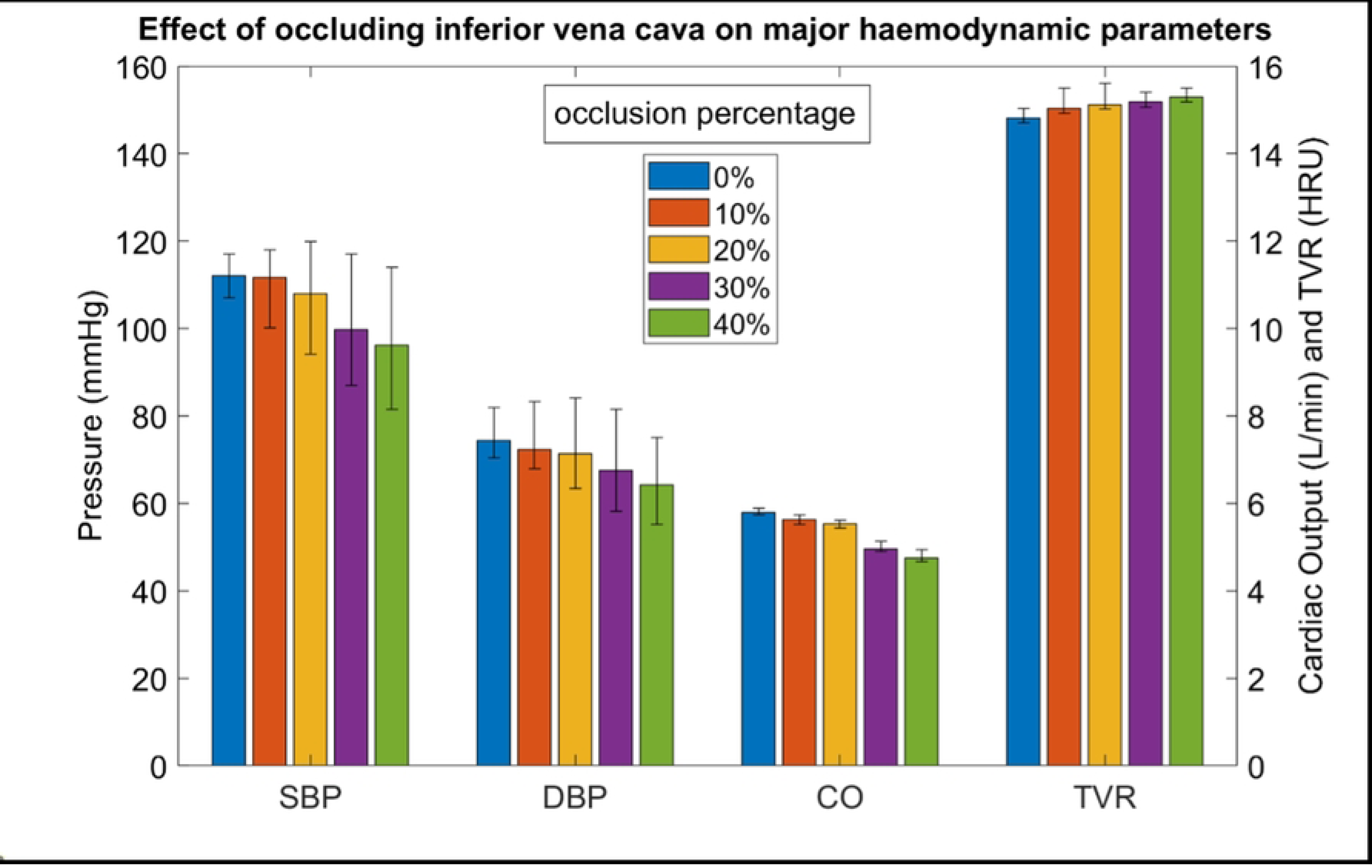
Effect of occluding the inferior vena cava on sBP, dBP, CO, and TVR using data from mathematical modelling. Error bars show the median values for each occlusion percentage and the interquartile range. TVR is shown in HRU/Woods units which is equivalent to mmHg*min/L.

### Scoping Review and Meta-Analysis

Overall, 79 studies were included in the scoping review (Supplementary file 2), 47 were case reports; the mean quality score for each case report was 3.12 out of 6. Items which had higher scores were ascertainment of exposure (which was the use of the prone position during pregnancy), length of follow-up, and detail of reporting. Domains which scored lowest included selection, causality, and reporting. These domains covered the presentation of a clear case selection method, explanation of alternative causes for observations, and adequate ascertainment of outcome (which was defined as livebirth and lack of fetal distress during proning). The selection method was unclear in all but two case reports. Lack of adequate ascertainment of outcome was usually due to insufficient duration of follow-up of birth outcomes, or due to lack of detail when reporting the outcome of fetal monitoring or livebirth. Case reports of women with COVID-19 infection were generally followed up for a shorter duration than their non-COVID-19 counterparts. In addition, most surgical case reports failed to report birthing outcomes or if fetal monitoring was conducted during the procedure.

Anecdotal reports of the prone position adopted during pregnancy accounted for over half of the evidence identified in this review, comprising a total of 75 individual cases. An overview of these case reports is shown in Supplementary file 3, including a summary of the method, monitoring and duration of proning, as well as the observed effect on various maternal and fetal outcome variables. The most common circumstance in which maternal proning was required was non-obstetric surgery, specifically that which required a posterior approach, such as brain or spinal surgery. The reported duration of prone positioning ranged between 1 and 6.5 hours. A surgical prone position was mostly achieved using variations of a traditional spinal operating frame. This generally comprised of sets of padded bolsters which were strategically placed to support the chest and hips whilst simultaneously freeing the pregnant abdomen. Care was taken to avoid external pressure on the gravid uterus as much as possible, as it was thought the resulting aortocaval compression may lead to fetal compromise.

Gestational age at the time of surgical prone positioning varied greatly, between 8 and 34 weeks’ gestation. The maximum gestation for adoption of the full prone position was 32 weeks’ with two additional cases of the modified three-quarter prone and the semi-prone positions utilised at 32 and 34 weeks’ gestation, respectively. Surgical prone positioning was mainly limited to the first and second trimesters due to the size of the gravid uterus in advanced pregnancy. As pregnancy progressed, delivery was commonly considered prior to surgical intervention, in order to avoid the logistical difficulties of performing prone surgery in advanced pregnancy.

The second most common indication for the maternal prone position was prone ventilation. Chest trauma, influenza infection, and COVID-19 infection were among the reported causes of respiratory distress which indicated the use prone ventilation in pregnant women. The duration of proning required per day ranged between 8 and 18 hours; often proning was required on consecutive days for above 16 hours at a time. As with spinal surgery, prone positioning was most commonly achieved using padded supports at the chest and hips which then allowed room for the pregnant abdomen. Notably, prone positioning was generally very successful in treating maternal refractory hypoxaemia, and in one case peripheral oxygen saturation spO_2_ was reportedly improved from 83% to 93% after just 30 minutes of prone ventilation. Regarding gestational age, cases of prone ventilation during pregnancy were generally limited to the first and second trimesters, with the exception of one case at 34 weeks’ gestation. After 34 weeks’ gestation, caesarean section was usually performed prior to treatment.

Maternal and fetal outcomes within the included case reports of prone positioning during pregnancy were generally very favourable. No maternal deaths were reported. One instance of maternal hypotension was reported during surgical prone positioning (sBP fell over 20% from baseline into a range of 90 to 100 mmHg) which persisted despite intervention for the duration of the five-hour surgery.

Livebirths were almost always reported, although some data on birth outcome was not available from individual case reports due to insufficient follow-up (22/75 cases). Two cases did not end in live birth, one case of spontaneous miscarriage that was discovered on ultrasound following surgical prone positioning of a twin pregnancy (although it is not clear when exactly during the course of the pregnancy this occurred) and one instance of termination of pregnancy which was performed following surgery in early pregnancy. No cases reported evidence of fetal distress during maternal proning, although prior to 24 weeks’ gestation fetal monitoring was often not deemed necessary due to lack of fetal viability. Gestational age at birth varied greatly, from 25 weeks 4 days to 40 weeks’ gestation, although birth outcome data was not always available. Preterm birth was relatively common in the included case reports (13/53, 24%), although this was more likely to be related to the critical illness experienced during these pregnancies than the prone position itself.

Characteristics of included larger-scale interventional studies are shown in Table 5. Meta-analysis of the three studies of maternal haemodynamic indices suggested an increase in sBP, but not dBP, though both analyses showed significant heterogeneity (I^2^ 79.8 and 76.2% respectively). Maternal HR increased in prone position. There were no changes in RR, oxygen saturation or fetal HR (Figure 8).

**Figure 8:**
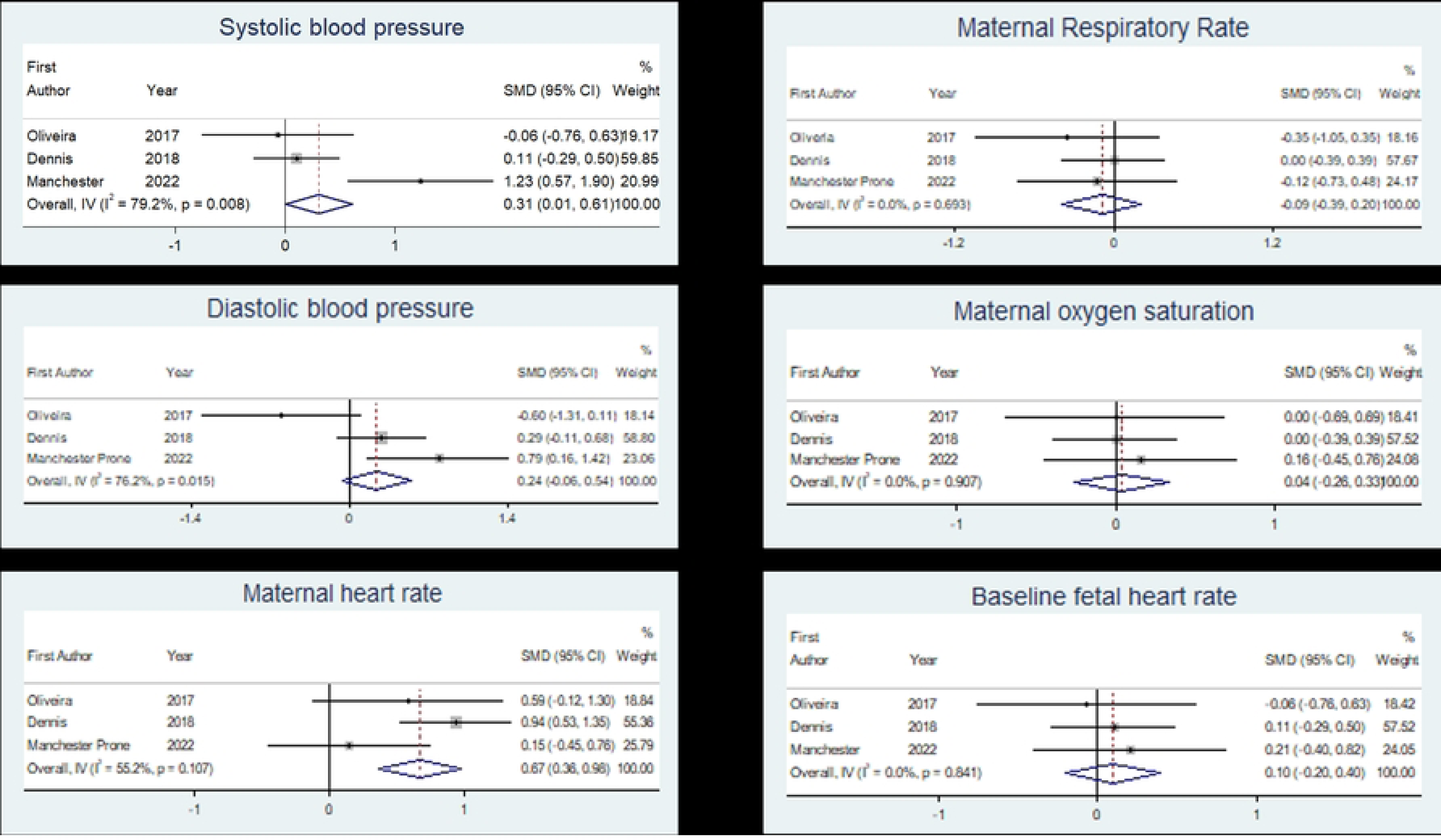
Forest plots of maternal haemodynamic characteristics and baseline fetal heart rate. Bold line demonstrates line of no effect. Blue diamond is the pooled effect with 95% confidence intervals.

**Table 5.** Characteristics of included studies for the meta-analysis of observational and interventional studies of altered maternal position.

| ***First Author/***<br>***Year/ Country*** | ***Study Type*** | ***Study***<br>***population*** | ***Data***<br>***extracted*** | ***Exposure*** | ***Summary of***<br>***Findings*** |
| --- | --- | --- | --- | --- | --- |
| Dennis<br>2018<br>Australia (10) | Prospective<br>observational<br>study | 50 healthy term<br>pregnant<br>women and 15<br>women with<br>preeclampsia. | sSBP, dBP,<br>MHR,<br>SpO<sub>2</sub>, RR,<br>FHR and<br>comfort<br>levels. | Data obtained in<br>Left Lateral and<br>Prone Position<br>(using specialised<br>pillow) after 5<br>minutes rest in<br>each position. | No change in<br>healthy women.<br>sBP reduced in<br>prone position in<br>pre-eclamptic<br>women, and in<br>33% of these<br>women it was by<br>over 10mmHg.<br>Around half of<br>women preferred<br>prone position. |
| Oliveira<br>2017<br>Brazil (9) | Randomised<br>controlled<br>trial | 33 healthy<br>pregnant<br>women | MHR,<br>SpO<sub>2</sub>, sBP,<br>dBP, RR,<br>FHR,<br>comfort. | Two different<br>sequences of<br>positions held for 6<br>minutes each.<br>Sequence 1:<br>Fowler's position,<br>Prone Position,<br>Supine Position,<br>Left Lateral,<br>Fowler's position<br>and repeat.<br>Sequence 2:<br>Fowler's position,<br>Prone Position,<br>Left Lateral, Supine<br>Position, Fowler's<br>position. Prone<br>position held using<br>specialised<br>stretcher. | sBP and RR were<br>decreased in<br>prone position<br>compared to left<br>lateral and a<br>decrease in DBP<br>and increase in<br>SpO<sub>2</sub> in prone<br>position<br>compared to<br>other positions.<br>All parameters<br>were in normal<br>limits, all women<br>report comfort in<br>all positions. |
| Current study<br>2023<br>United<br>Kingdom | Prospective<br>observational<br>study | 21 healthy<br>pregnant<br>women | sBP, dBP,<br>MHR,<br>SpO<sub>2</sub>, RR,<br>FHR<br>comfort. | Left lateral<br>position for 20<br>minutes, Prone<br>position for 30<br>minutes, returned<br>to left lateral<br>position for 20<br>minutes using a<br>supportive<br>cushion. | sBP, dBP<br>increased in<br>prone position.<br>No change in<br>MHR, RR or FHR. |

sBP = Systolic Blood Pressure, dBP = Diastolic Blood Pressure, MHR = Maternal Heart Rate, SpO<sub>2</sub> = Oxygen Saturation, RR = Respiratory Rate, FHR = Fetal Heart Rate

## Discussion

Prone position was associated with a reduction in maternal CO, largely attributable to reduced SV. The mechanism behind this remains uncertain, but could be a result of reduced venous return. This could be due to upward compression from the Anna cushion on the inferior vena cava or gravitational pooling of blood in the uteroplacental circulation. Alternatively, it could be due to thoracic compression in prone position; this has been postulated to increase left ventricular resistance, thereby reducing SV (18,19). Maternal haemodynamics returned to baseline when moved back into left lateral, indicating a transient rather than lasting positional effect. The scoping review found minimal clinical adverse effects of maternal prone positioning at various stages of pregnancy.

If the decline in venous return were due to upward compression of the inferior vena cava, one would expect a decline in both CO and BP, as seen in supine hypotensive syndrome. The computational model predicted this trend, however, the patient measurements only demonstrated a decrease in CO; instead there was an increase in BP in prone position. This could in part be due to auto-regulatory mechanisms in the body. However, this discrepancy could also be attributed to the effect of gravity. The gravitational effect on BP with respect to body position is well understood (20). BP increases/decreases by approximately 7 mmHg for every 10 cm distance below/above the heart. As the difference between the heart and the right upper arm is approximately 20 cm, it would be expected that the BP recording in the left lateral position would be approximately 15 mmHg lower than what was measured in the prone position, from the effect of gravity alone. It is also important to remember that the brachial cuff BP is actually attempting to estimate the pressure in the aorta; this is why the protocol of cuff BP measurement requires the patient to have their arm supported at heart level (it is typically raised and rested on a table). This is also particularly important when other parameters are calculated, such as TVR, which is typically calculated as MAP=TVR*CO (as mean venous pressure is often wrongly assumed to be 0 mmHg). MAP and CO should be taken at the same location in the arterial system, and as the CO measurement is restricted to the aorta (by virtue of the fact flow splits into other blood vessels), the BP also needs to be measured/estimated at the aorta to be a reliable measure of resistance. This could explain the discrepancies in the patient measurement trends, as the aortic BP estimates from the brachial cuff measure will be approximately 15 mmHg lower than the actual aortic pressure, and this means the TVR calculation will not be reliable in the left lateral position as it is being calculated using an incorrect pressure estimate of the aorta. If 15 mmHg is added to the left lateral position estimate of aortic BP, the expected and the model predicted trend, would be observed. In this way, prone position could actually reduce aortic pressure when compared to left lateral.

The inconsistency of the model predictions across the group is likely due to a lack of information on pressures in the systemic veins, pulmonary system arteries and veins, and autoregulatory mechanisms in the body. Not everyone is affected equally in the prone position with not everyone developing supine hypotensive syndrome. Some of the individual variation might be due to the blood being redirected to the heart alternatively via the azygos vein. The patients that develop supine hypotensive syndrome tend to have less flow increase in the azygos vein compared to those individuals that remain normotensive (21); the location of compression along the inferior vena cava plays a key role in this.

A similar decline in SV and increase in vascular resistance have been demonstrated in previous non-pregnant prone positioning studies (18,19). On the other hand, our findings differed from previous studies in pregnancy (9,10), which did not demonstrate an increase in maternal BP. As explained above, this could be attributed to varying cuff positions and therefore gravitational effects on BP. Alternatively, the cardiovascular regulatory mechanisms from neural, renal and endocrine systems impact on maternal haemodynamics within different timeframes. It is therefore plausible that differences seen between studies reflect different phases of cardiovascular adjustment due to varying protocol timings (18). Previous maternal studies only maintained position changes for 5 to 6 minutes (9,10), compared with 30 minutes in this study.

The impact of these positional haemodynamic changes on fetal wellbeing remain uncertain. Although fetal HR, decelerations and variability were unaltered by maternal position, accelerations were reduced in prone position and one woman had a fetal bradycardia. On review of this woman’s simultaneous haemodynamic measures there appeared to be no temporal relationship between maternal haemodynamic alterations and her fetal bradycardia. This suggests that this was unlikely due to prone-induced altered maternal haemodynamics. The significance of reduced fetal accelerations in prone position is uncertain.

To our knowledge, this is the first study to simultaneously investigate the maternal cardiorespiratory effects of prolonged prone positioning in late pregnancy, whilst continuously monitoring fetal HR. The multimodal approach combined with a scoping review adds strength to this evaluation of the clinical consequences of prone positioning. Finally, the scoping review included all available evidence, recognising that the clinical evidence is mostly individual cases. The main limitation of this study is the comparatively small sample size, making it difficult to determine whether the witnessed bradycardia was a random occurrence or a consequence of prone positioning. Limitations of the scoping review were that high quality studies evaluating the effects of prone positioning were scarce and a significant proportion of the case reports omitted information about maternal or fetal outcomes. There is also the possibility that there is a publication bias towards healthy outcomes for mother and baby. In addition, there was a wide variation in gestation in this cohort. Further work is needed to explore any differences in positional haemodynamics at varying gestations. This could facilitate exploration into the impact of differing abdominal girth and uteroplacental circulation on maternal haemodynamics, thereby providing insight into the mechanism linking prone position and altered maternal haemodynamics. Furthermore, development of the cushion, in terms of shape and consistency, could provide insight into the effect of varying degrees of vena caval compression on maternal haemodynamics. If vena caval compression is the key mechanism linking prone position and reduced maternal CO, further development of the cushion has the potential to optimise prone position safety for mother and fetus.

## Conclusion

Prone position was associated with a reduction in CO, however the impact on fetal wellbeing remains uncertain. It is therefore unclear from our data whether extended periods of prone positioning of up to 30 minutes (i.e. during physical therapy) are safe in late pregnancy. Although comfort was not altered from the start to finish of the protocol, anxiety scores were marginally reduced. Further work is needed to optimise the safety of prone positioning in pregnancy.

## Data Availability

The data underlying the results presented in the study are available upon request from Laura Ormesher

## Acknowledgments

This study was funded by the Dowager Countess Eleanor Peel Medical Research Trust. The authors are grateful to Professor Peter Stone and Professor Jane Warland for their advice and expertise during the planning, conduct and analysis of this study.

## Author contributions

AH and KB conceived the overall study and applied for funding. JC, LP, HP and AH recruited participants and conducted the experimental protocol. LO and LW undertook the fetal behavioural assessment. LO and AH undertook the statistical analysis. JC, AF-H, DH and AH conducted the scoping review. CP, JMC, RvL undertook the mathematical modelling. LO produced the first draft of the manuscript; all authors contributed to the submitted version.

